# Greater Stress Response Network (SRN) Connectivity is Associated with Higher Worry in Men and Women Especially in Later Life

**DOI:** 10.64898/2026.11.05.26367375

**Authors:** Emma Hudson, Christopher Chae, Dana L. Tudorascu, Meryl A. Butters, Rebecca C. Thurston, Carmen Andreescu, Helmet T. Karim

## Abstract

**Background:** Late-life anxiety is common and disabling, with pathological worry as a core feature. The Stress Response Network (SRN: amygdala, bed nucleus of the stria terminalis, paraventricular nucleus), which modulates threat and stress reactivity, and its interaction with the Executive Control Network (ECN), anchored by the dorsolateral prefrontal cortex, which supports emotional regulation via top-down cognitive control, may underlie individual differences in worry severity. We hypothesize higher SRN connectivity to be associated with greater worry severity and that this relationship may be moderated positively by older age, and female sex.

**Methods:** We recruited older adults with varying worry. Resting-state fMRI was used to compute SRN connectivity and SRN-ECN connectivity. Regression models tested associations between connectivity and worry (PSWQ), anxiety (HARS), and rumination (RSQ), adjusting for age, sex, race, and education. Exploratory models examined age and sex moderation and associations with emotion regulation/suppression and cognitive function.

**Results:** Among 131 participants who were ≥ 50 years old, with varying levels of worry severity, SRN connectivity was significantly associated with greater worry severity (B=23.51 (SE = 12.12), p<0.05), whereas SRN-ECN connectivity was not. SRN connectivity was lower with greater age (B=-0.003 (SE=0.001), p=0.04), yet older individuals reported greater worry than younger with comparable connectivity. Greater language domain cognitive function was associated with lower SRN connectivity but higher SRN-ECN connectivity, while greater overall cognitive function was associated with lower SRN-ECN connectivity. Women reported lower use of daily expressive suppression and higher performance across cognitive measures than men. Higher daily use of expressive suppression was not significantly associated with either connectivity measure.

**Discussion:** SRN connectivity, rather than SRN-ECN connectivity, may serve as the primary neural correlate of worry, shaped by age and cognitive function.

## Introduction

Anxiety disorders are among the most prevalent psychiatric conditions in the United States, with lifetime prevalence estimates at 33%.^1^ Anxiety symptoms that do not fall under a categorical diagnosis are prevalent in community samples, up to 52%^2^, and are also associated with high comorbidity with other mental health disorders.^3^ The burden of late-life anxiety is particularly concerning as it is associated with increased morbidity and mortality, including high vascular risk and accelerated cognitive decline^4^, and has been noted as one of the top five contributors to years lived with disability in the US.^5^ Anxiety has a second peak of incidence starting after age 50 years^6,7^ and women are 1.7-3 times more likely than men to experience anxiety in late life.^8–10^ A core defining feature of generalized anxiety disorder^11^ is pathological worry^12,13^, characterized as a persistent cognitive process marked with negative affect and poor inhibitory control of excessive and pervasive negative thoughts about future events.

Research suggests that the neural pathways of anxiety and worry create a hypervigilant state which resembles a chronic stress response with enhanced physiological activation of cardiovascular, endocrinological, immunological, and neurovisceral activity.^14^ Thus, we aim to explore the neural circuitry engaged in chronic stress response. The Stress Response Network (SRN) encompasses regions such as the Paraventricular Nucleus (PVN), which plays a key role as an autonomic control center initiating hypothalamic-pituitary-adrenal (HPA) axis responses^15,16^; the extended amygdala, which is critical for assigning emotional salience and integrating cortical and sensory input^17,18^; the Bed Nucleus of the Stria Terminalis (BNST), which is involved in fear responses and coordinating HPA output^19^; and the habenula, whose role in anxiety has gained increasing interest due to its connections with emotional processing, reward, and decision making.^20^

These regions have each been consistently linked to anxiety: the amygdala and BNST with modulating anxiety^21,22^; the PVN as a key regulator of anxiety;^23^ and the habenula as a critical target of stress effects^24^. While the amygdala and BNST are central to fear and anxiety processing, the PVN and habenula play pivotal roles in modulating stress responses. Emerging research also highlights the involvement of the PVN and BNST in emotion reactivity and regulation related to anxiety and worry in adults over age 50^25–27^. Greater PVN connectivity with cortical regulatory regions, such as the subgenual anterior cingulate and parahippocampal cortex, has been linked to greater worry, whereas amygdala connectivity with temporal regions shows an association with lower anxiety and perceived stress^28^. Consistent with its role in anticipatory threat processing and prolonged fear states^29^, the BNST also shows heightened connectivity in individuals with higher anxiety, underscoring its role in sustained worry^30^.

The Executive Control Network (ECN) is central to emotional regulation and cognitive control, but its functioning is often compromised in anxiety and stress-related disorders. Normally, prefrontal regions exert top-down inhibition over the amygdala, a mechanism weakened in anxiety^31^. Through connections linking the dorsal anterior cingulate (dACC) and dorsomedial prefrontal cortex (dmPFC) to the amygdala, the ECN closely interacts with the SRN to regulate emotional reactivity^32^. In healthy states, stronger ECN–SRN coupling supports adaptive cognitive control during stress^33,34^. In anxiety disorders, however, this circuitry becomes dysregulated, with aberrant connectivity contributing to persistent worry and impaired emotion regulation^35,36^. Thus, ECN–SRN interactions represent a critical target for understanding the neural mechanisms of anxiety regulation.

Elevated worry^37^ and anxiety^38^ are more common in women than men, though the sex difference in worry decreases with age^8^. Older adults’ greater use of cognitive emotion-regulation strategies^39^ and sex differences in effortful control processes^40,41^ may partly explain this pattern. Research finds men to show stronger amygdala-prefrontal connectivity than women when processing negative emotion, an effect associated with testosterone levels and more action-oriented response to threat^42^, indicating sex-specific SRN–ECN dynamics. Hormonal factors in women may also partially explain these differences: in premenopausal women, higher levels of specific circulating estrogen metabolites were associated with higher connectivity within the default mode and sensorimotor networks, whereas in postmenopausal women, higher circulating estradiol was associated with higher locus coeruleus-executive control network connectivity^43^. Although these associations were measured in plasma and stool rather than brain tissue, this suggests that peripheral estrogen levels may relate to central network connectivity, though the extent to which they reflect direct estrogenic action in the brain remains unclear. Furthermore, estrogen decline during the menopausal transition has been linked to disruption in estrogen-regulated systems, affecting multiple domains of cognitive function, including reduced top-down cortical control, among other neurologic dysfunctions^44^.

Separately, aging more broadly, independent of menopausal status, has been associated with reduced network segregation, reflecting an age-related shift toward greater between-network relative to within-network connectivity^45^. However, this age-related reorganization also appears to differ by sex, where men show relatively stable inter-network connectivity across the lifespan, while women show greater age-related change in connectivity^46^. Together, these age- and sex-related patterns in network connectivity underscore the importance of examining SRN-ECN interactions specifically in later life.

Overall, research indicates that connectivity patterns within the SRN and between the SRN and ECN may help explain differences in worry severity, with this relationship further shaped by sex and age-related variation. In this study, we aim to investigate exactly that and examine SRN and SRN-ECN connectivity and their associations with age, sex, and worry. We hypothesized that higher SRN connectivity would be associated with greater worry severity, and that this relationship would be moderated by age such that the associations between SRN connectivity and worry would be stronger among older adults and moderated by sex such that SRN connectivity and worry would be stronger in women than men.

## Methods

### Participants and Study Design

We recruited participants into the Functional neuroanatomy correlates of worry in older adults (FINA) study (R01 MH108509). They were recruited through a variety of sources including a website resource from the university, recommendations, flyers, and ads. This study was approved by the University of Pittsburgh Institutional Review Board. All participants gave written informed consent prior to participating in the study.

We recruited 131 participants who were ≥ 50 years old, with varying levels of worry severity. We recruited participants to have a normal distribution on worry severity in the sample as measured by the Penn State Worry Questionnaire (PSWQ)^38^. The presence of a categorial mood or anxiety disorder (diagnosed by the Structured Clinical Interview for DSMV SCID V REF) was allowed but not required. Participants were excluded if they were diagnosed with autism spectrum disorders, intellectual development disorder, or any form of psychosis or bipolar disorder. Other exclusion criteria were: a diagnosis of major neurocognitive disorder (e.g., dementia), a modified mini-mental state exam (3MSE) score < 84, a diagnosis of personality disorder, high suicide risk, use of antidepressants within the last five to fourteen days, history of drug/alcohol abuse within last six months, use of high doses of benzodiazepines (greater than equivalent to 2mg/day of lorazepam), uncorrected vision problems that would preclude neuropsychiatric testing, below 6^th^ grade level of reading, clinical diagnosis of cerebrovascular accident, multiple sclerosis, vasculitis or significant head trauma. Participants with ferromagnetic objects in body, claustrophobia, or too large to fit in an MR scanner were also excluded. Some participants underwent washout of antidepressants when appropriate [e.g. poor response to current antidepressants, determined after evaluation by a geriatric psychiatrist (CA)]. Participants on low dose psychotropics for other medical conditions (e.g. peripheral neuropathy) were allowed to continue in most cases (n=5). Two participants were tapered off psychotropics.

### Assessments

We collected demographic data (age, sex, race, education). We assessed worry severity with the Penn-State Worry Questionnaire (PSWQ)^47^, global anxiety with the Hamilton Anxiety Rating Scale (HARS)^48^, and rumination with the Response Styles Questionnaire Rumination Subscale (RSQ)^49^. We additionally assessed depression with the Montgomery-Asberg Depression Rating Scale (MADRS)^50^, neuroticism with the subscale from the Five Factor Inventory (NEO-FFI)^51^, stress with the Perceived Stress Scale (PSS)^52^, habitual use of cognitive reappraisal and suppression with the Emotion Regulation Questionnaire (ERQ)^53^, overall illness burden with the cumulative illness rating scale for geriatrics (CIRSG)^54^, and general cognitive function with the 3MSE^55^.

We also assessed cognitive function using a comprehensive battery that included the Repeatable Battery for the Assessment of Neuropsychological Status (RBANS) and executive function (EF) with select subtests from the Delis-Kaplan Executive Function System (DKEFS^56^. Specifically, the scaled trail making conditions 4 v. 5 (set shifting) and scaled color-word condition 3 (inhibition) were z-scored and combined to compute a measure of executive function. Regular meetings with an expert neuropsychologist (MAB) allowed for the adjudication of cognitive status and diagnosis of each subject.

### MRI Acquisition

We acquired MR data at the MR Research Center at the University of Pittsburgh using a 3T Siemens MAGNETOM Prisma scanner and a 32-channel head coil. We collected a sagittal whole-brain T1-weighted magnetization prepared rapid gradient echo (MPRAGE) with repetition time (TR)=2400ms, echo time (TE)=2.22ms, flip angle (FA)=8deg, field of view (FOV)=320x300 with 208 slices, 0.8mm^3^ isotropic resolution, 0.4mm slice gap, and GeneRalized Autocalibrating Partial Parallel Acquisition (GRAPPA) with acceleration factor of 2 (total time 6.63min). A sagittal, whole-brain T2-weighted Sampling Perfection with Application optimized Contrasts using different flip angle Evolution (SPACE) was collected with TR=3200ms, TE=563ms, FA=120deg, FOV=320x300 with 208 slices, 0.8mm^3^ isotropic resolution, no slice gap, and GRAPPA with acceleration factor of 2 (total time 5.95min). An axial, whole-brain T2-weighted Fluid Attenuated Inversion Recovery (FLAIR) was collected with TR=10,000ms, TE=91ms, FA=135deg, inversion time (TI)=2,500ms, FOV=320x320 with 104 slices, 0.8mm x 0.8mm x 1.6mm resolution, no slice gap, and GRAPPA with acceleration factor of 2 (total time 5.95min). Participants were in the MR scanner for approximately 45-60 minutes as we also collected functional MRI as well. The entire scan lasted approximately an hour. Resting state data was acquired over a 6-minute interval while participants were directed to view a white crosshair in the center of a black screen and told to stay awake. Whole-brain T2*-weighted BOLD images were acquired axially using gradient echo-planar imaging (EPI) sequence with the following parameters: repetition time (TR)/echo time (TE)=1000/30ms, flip angle (FA) 45°, FOV 96 × 96 with 60 axial slices, 2.3mm^3^ isotropic resolution with 2.3 mm slice gap, and multiband acceleration factor of 5.

### Structural Processing

All processing was done in statistical parametric mapping toolbox^57^ (SPM12) in MatLab 2018b (MathWorks, Natick, MA). Both T2-SPACE and FLAIR were registered to the MPRAGE and then were all input into a multispectral segmentation that bias corrects each image and segments them into gray matter, white matter, cerebrospinal fluid, skull, soft-tissue, and air^58^. We adjusted the number of Gaussians to two to more accurately identify white matter. We generated an intracranial volume mask by thresholding gray/white matter and cerebrospinal fluid probability maps by 0.1, then conducted image filling and closing to generate a final mask. This was used to skull strip the MPRAGE.

### Resting State Processing

The resting state fMRI was processed with SPM12 in MatLab and the FMRIB Software Library v6.0 Brain Extraction Tool (FSL BET) ^59^. Images were slice-time corrected and motion-corrected to the mean functional image with a rigid body transformation. The functional images were skull-stripped using BET, visually inspected, and coregistered to the MPRAGE before being transformed to MNI space using the deformation field generated in structural processing. The images were spatially smoothed with an 8 mm Gaussian kernel. The following were regressed out of each voxel: six rigid-body motion parameters, first five principal components of the white matter and cerebral spinal fluid, and sinusoids corresponding to unwanted frequencies outside of the resting state band 0.008 and 0.15 Hz of interest for resting state analysis (i.e., a band-pass filter). By doing this in one step, we do not reintroduce artifact/noise into our signal^60^. We used the ArtRepair toolbox^61^ to calculate several subject-level parameters of motion including max translations, average root mean squared, average scan-to-scan motion, and percent of headjerks (framewise displacement >0.5 mm).

We defined the SRN as the left/right amygdala (from AAL3 atlas), while the following were defined based on a previously defined probabilistic atlas^28,62,63^: ventral BNST, dorsal BNST, and PVN using a threshold of 0.2. The ECN was defined by seeding the left dlPFC (MNI -38, 44, 26) in neurosynth^64^, thresholding that map at 0.15, and then dividing them into regions using the AAL3 atlas resulting in a total of 36 regions. We extracted the principal time series from each region and then computed an all to all connectivity matrix (40 x 40 regions). We extracted the SRN connectivity as the average connectivity from all 4 regions to all other 4 regions (lower triangle) and the ECN-SRN connectivity as the average connectivity between all 4 SRN regions to all 36 ECN regions.

### Statistical Analysis

We conducted independent t-tests for continuous variables and chi-square tests for categorical variables. We conducted three regression analyses with each of the following dependent variables worry (PSWQ), anxiety (HARS), and rumination (RSQ) with the following independent variables age, sex, race, education, SRN connectivity, and SRN-ECN connectivity. We conducted analyses with HARS and RSQ as they are commonly considered phenotypes of generalized anxiety disorder (GAD).

We found that only SRN connectivity correlated with PSWQ (see results), so we conducted two exploratory analyses. First, we sought to see whether SRN connectivity was correlated with age and sex, so we used SRN connectivity as a dependent variable and age, sex, race, and education as independent variables. We then conducted an exploratory analysis to evaluate whether there was a three-way interaction with PSWQ by regressing the SRN connectivity as a dependent variable onto age x sex x PSWQ (with lower order second and third order interactions) while adjusting for education and race. Finally, we evaluated whether SRN or SRN-ECN connectivity were associated with ERQ suppression and cognitive function while adjusting for age, sex, race, and education. For all models, we evaluated the variance inflation factor (VIF) and ensured they were less than 5. We evaluated residuals for normality. We evaluated residuals plots to check for regression assumptions. All analyses were conducted in R version 4.4.2. We generated table 1 with the table1 package in R.

**Table 1.** Demographic and clinical characteristics comparing men and women.

|  | **Male** | **Female** | **P-value** |
|---|---|---|---|
|  | (N=50) | (N=81) |  |
| Age | 61.9 (9.25) | 60.8 (8.17) | 0.501 |
| Education (years) | 16.0 (2.44) | 15.8 (2.36) | 0.708 |
| Race (White) | 43 (86.0%) | 63 (77.8%) | 0.35 |
| CIRSG | 4.04 (3.36) | 4.71 (3.57) | 0.296 |
| PSWQ | 49.5 (14.6) | 51.6 (14.7) | 0.418 |
| HARS | 8.77 (7.85) | 9.54 (6.60) | 0.572 |
| RSQ | 37.0 (12.5) | 39.8 (13.2) | 0.237 |
| MADRS | 8.00 (8.32) | 8.55 (7.75) | 0.715 |
| Neuroticism | 20.1 (9.85) | 20.9 (10.8) | 0.686 |
| PSS | 19.1 (5.35) | 20.4 (5.59) | 0.188 |
| Reappraisal | 28.8 (7.70) | 29.9 (7.18) | 0.45 |
| **Suppression** | **16.0 (5.06)** | **12.8 (5.17)** | **<0.001** |
| **Immediate Memory Index Score** | **95.0 (18.8)** | **106 (14.8)** | **<0.001** |
| Visuospatial Index Score | 100 (16.1) | 98.9 (17.4) | 0.713 |
| **Language Index Score** | **97.6 (9.62)** | **104 (10.5)** | **<0.001** |
| Attention Index Score | 104 (15.6) | 108 (14.9) | 0.18 |
| **Delayed Memory Index Score** | **97.1 (15.9)** | **106 (14.5)** | **<0.005** |
| **Total Cognitive** | **98.3 (13.1)** | **106 (14.8)** | **<0.005** |
| SRN connectivity | 0.497 (0.133) | 0.525 (0.112) | 0.224 |
| **SRN-x-ECN connectivity** | **0.0434 (0.0955)** | **0.0762 (0.100)** | **0.0641** |

## Results

We found that compared to men, women showed lower habitual use of suppression (ERQ) for emotion regulation, greater immediate and delayed memory, greater language cognitive function (fluent use of expressive and receptive language), and greater total cognitive function (see table 1).

We found that greater SRN connectivity was associated with greater worry severity (table 2, figure 1A). We found no association between SRN or SRN-ECN connectivity and either HARS or RSQ (not shown).

**Figure 1.**
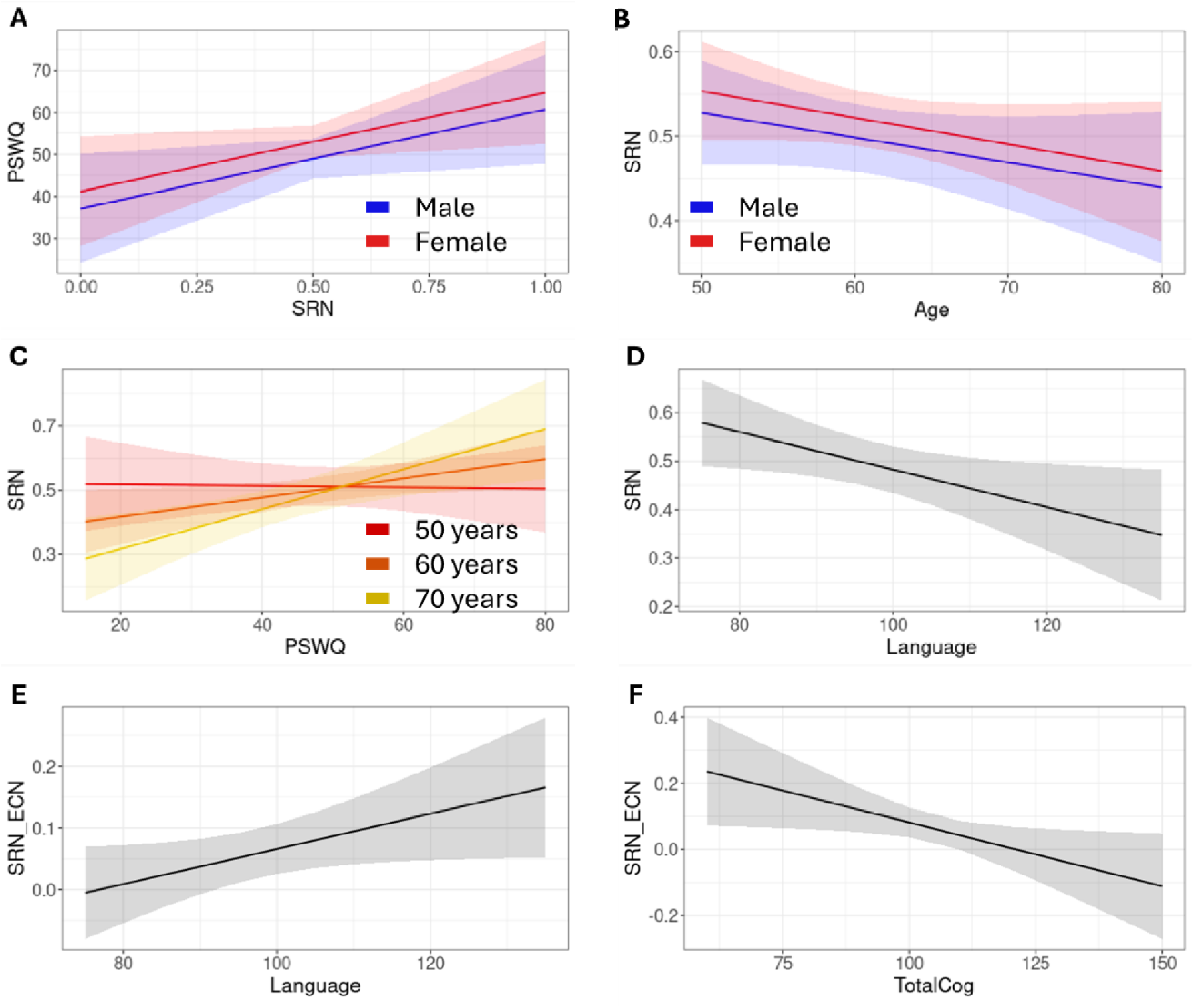
Associations between age, PSWQ, and SRN connectivity. (A) Greater SRN connectivity was associated with PSWQ (this was true for men and women). (B) Greate age was associated with lower SRN connectivity in both men and women. (C) The SRN by PSWQ association is moderated by age, wherein older individuals with higher SRN have greater PSWQ. The different lines show the fifth, sixth, and seventh decade of life as predicted values from the model. Greater language function is associated with lower SRN connectivity (D) but greater SRN-ECN connectivity (E). Greater total cognitive function is associated with lower SRN-ECN connectivity.

**Table 2.** Association between PSWQ and age, sex, race, education, SRN, and SRN-ECN connectivity as independent variables.

| **Variable** | **B (SE)** | **ß** | **t-statistic** | **p-value** |
|---|---|---|---|---|
| Intercept | 70.63 (16.04) | NA | 4.4 | <0.001 |
| Age | -0.34 (0.18) | -0.18 | -1.9 | 0.0647 |
| Sex (Ref M) | 4.02 (2.95) | 0.13 | 1.4 | 0.1765 |
| Race (Ref White) | -6.94 (3.82) | -0.18 | -1.8 | 0.0727 |
| Education | -0.74 (0.65) | -0.12 | -1.1 | 0.2597 |
| **SRN connectivity** | **23.51 (12.12)** | **0.19** | **2.0** | **<0.05** |
| SRN-x-ECN connectivity | -17.28 (14.91) | -0.12 | -1.2 | 0.2494 |

We conducted two sets of exploratory analyses. We found that greater age was associated with lower SRN connectivity (table 3, figure 1B). We found that the association between SRN connectivity and worry severity was significantly moderated by age, wherein older individuals with higher SRN connectivity showed greater worry regardless of sex (table 4, figure 1C).

**Table 3.** Exploratory regression between SRN connectivity and the following independent variables age, sex, race, and education.

| **Variable** | **B (SE)** | **ß** | **t-statistic** | **p-value** |
|---|---|---|---|---|
| Intercept | 0.56 (0.12) | NA | 4.63 | <0.001 |
| **Age** | **-0.003 (0.001)** | **-0.20** | **-2.07** | **0.04** |
| Sex (Ref M) | 0.02 (0.02) | 0.09 | 0.95 | 0.34 |
| Race (Ref White) | 0.01 (0.03) | 0.03 | 0.32 | 0.75 |
| Education | 0.008 (0.005) | 0.15 | 1.49 | 0.14 |

**Table 4.** Exploratory analysis exploring three-way interaction with SRN connectivity as dependent variable and the following interaction with lower order interaction terms: age x sex x PSWQ while adjusting for education and race.

| **Variable** | **B (SE)** | **ß** | **t-statistic** | **p-value** |
|---|---|---|---|---|
| Intercept | 0.42 (0.09) | NA | 4.67 | <0.001 |
| Age | -0.001 (0.02) | -0.01 | -0.07 | 0.95 |
| Sex (Ref M) | 0.01 (0.02) | 0.03 | 0.32 | 0.75 |
| **PSWQ** | **0.05 (0.02)** | **0.41** | **2.48** | **0.01** |
| Race (Ref White) | 0.02 (0.03) | 0.06 | 0.62 | 0.54 |
| Education | 0.01 (0.005) | 0.11 | 1.03 | 0.31 |
| Age x Sex (Ref M) | -0.02 (0.03) | -0.13 | -0.90 | 0.37 |
| **Age x PSWQ** | **0.04 (0.02)** | **0.31** | **2.17** | **0.03** |
| Sex (Ref M) x PSWQ | -0.04 (0.03) | -0.27 | -1.61 | 0.11 |
| Age x Sex (Ref M) x PSWQ | -0.02 (0.03) | -0.14 | -0.97 | 0.33 |

We found that greater language cognitive function was associated with lower SRN connectivity (table 5, figure 1D) but greater SRN-ECN connectivity (table 6, figure 1E). Greater total cognitive function was associated with lower SRN-ECN connectivity (table 6, figure 1F). Exploratory analyses were not moderated by sex.

**Table 5.** Exploratory analysis with SRN connectivity as dependent variable and the following independent variables age, sex, race, education, suppression, total cognitive function, delayed memory, immediate memory, and language function. These variables showed differences by sex.

| **Variable** | **B (SE)** | **ß** | **t-statistic** | **p-value** |
|---|---|---|---|---|
| Intercept | 0.84 (0.18) | NA | 4.59 | <0.001 |
| Age | - 0.0008 (0.002) | -0.05 | -0.47 | 0.64 |
| Sex (Ref M) | 0.06 (0.03) | 0.22 | 1.67 | 0.10 |
| Race (Ref White) | 0.02 (0.04) | 0.07 | 0.57 | 0.57 |
| Education | 0.006 (0.006) | 0.11 | 0.96 | 0.34 |
| Total Cognitive Function | 0.003 (0.002) | 0.37 | 1.55 | 0.13 |
| ERQ Suppression | - 0.002 (0.003) | -0.08 | -0.70 | 0.48 |
| **Language** | **- 0.004 (0.001)** | **-0.35** | **-2.28** | **0.03** |
| Immediate Memory | - 0.002 (0.001) | -0.22 | -1.33 | 0.19 |
| Delayed Memory | -0.002 (0.002) | -0.16 | -0.95 | 0.35 |

**Table 6.** Exploratory analysis with SRN-ECN connectivity as dependent variable and the following independent variables age, sex, race, education, suppression, total cognitive function, delayed memory, immediate memory, and language function. These variables showed differences by sex.

| Variable | B (SE) | ß | t-statistic | p-value |
|---|---|---|---|---|
| Intercept | 0.25 (0.14) | NA | 1.61 | 0.11 |
| Age | - 0.002 (0.002) | -0.13 | -1.14 | 0.26 |
| Sex (Ref M) | - 0.001 (0.03) | 0.00 | -0.03 | 0.98 |
| Race (Ref White) | - 0.01 (0.03) | -0.04 | -0.33 | 0.74 |
| Education | - 0.01 (0.005) | -0.21 | -1.86 | 0.07 |
| **Total Cognitive Function** | **- 0.003 (0.002)** | **-0.53** | **-2.21** | **0.03** |
| Suppression | - 0.002 (0.002) | -0.12 | -1.05 | 0.30 |
| **Language** | **0.003 (0.001)** | **0.30** | **2.00** | **0.05** |
| Immediate Memory | 0.001 (0.001) | 0.16 | 1.00 | 0.32 |
| Delayed Memory | 0.001 (0.001) | 0.14 | 0.82 | 0.42 |

## Discussion

Greater SRN connectivity was associated with more severe worry, and this relationship was stronger in older individuals despite the finding that SRN connectivity is lower with greater age. Compared to men, we found that women in this cohort showed lower habitual use of suppression for emotion regulation and higher cognitive performance, including memory, language, and overall cognition. Higher language function was linked to lower SRN connectivity but higher SRN–ECN connectivity, while overall cognitive function was associated with lower SRN–ECN connectivity. These findings suggest that in women, greater language cognitive function may support stronger ECN-SRN connectivity, reflecting greater reliance on top-down cognitive control over stress and worry-related neural circuits, potentially through the engagement of verbal or narrative-based strategies. Although women also reported lower habitual use of suppression than men, suppression itself was not significantly associated with SRN or SRN-ECN connectivity, suggesting that this sex difference is better explained by language-based cognitive control rather than reduced emotional suppression.

As hypothesized, higher SRN connectivity was associated with greater worry severity. This relationship was moderated by age, with older adults showing stronger associations between SRN connectivity and worry despite an overall age-related “decline” in SRN connectivity. This pattern suggests that although SRN connectivity is lower with greater age, higher connectivity in older individuals may reflect compensatory or less efficiently regulated activity, potentially due to age-related reductions in top-down cognitive control. The age-related SRN “declines” may partly explain findings that show that lower worry is associated with greater age, which may reflect a lowering in tonic SRN connectivity due to aging. The age-related “decrease” in SRN hyperactivity may reflect broader changes in emotion regulation and stress physiology across the lifespan. Aging is associated with altered HPA axis function, including reduced cortisol response to acute stress leading to reduced tonic stress signaling and attenuated SRN activation in older adults.^65,66^

Higher language function was linked to stronger SRN–ECN connectivity and lower SRN connectivity, consistent with the recruitment of cognitive control networks for adaptive regulation of stress and worry. In contrast, higher overall cognitive function was associated with lower SRN–ECN connectivity, potentially suggesting that individuals with stronger overall cognitive resources may rely less on SRN-ECN engagement, needing less compensatory recruitment of top-down control to manage stress. Together, these findings highlight that cognitive abilities, emotion regulation strategies, and network connectivity interact to shape individual differences in worry.

Functional connectivity within these networks is lower with greater age, accompanied by lower network segregation, cortical atrophy, and cognitive decline^45^. During midlife, women may experience more pronounced changes in functional connectivity potentially due to hormonal influences throughout the transition from perimenopause to post-menopause ^68^. Estradiol has been positively associated with connectivity involving the parahippocampus and other regions, while declining estradiol levels during menopause have been associated with reduced connectivity in the amygdala, hippocampus, and frontal regions^45,67,68^,. These age- and hormone-related changes may reduce the efficacy of top-down regulation and dampen stress responsiveness, explaining why older adults may show altered worry patterns despite lower overall SRN connectivity.

Overall, our findings suggest that sex, age, and cognitive function jointly shape SRN and SRN–ECN connectivity and their influence on worry. In women, higher language cognitive function appears to drive stronger ECN-SRN coupling, reflecting adaptive top-down regulation, rather than the lower emotional suppression women reported compared to men, which was not associated with connectivity in this sample. Age-related declines in network connectivity and cognitive control may weaken these mechanisms, leading to increased worry in older adults. This may be especially salient in older women due to age-related hormonal changes that occur after 50 years of age.

Several limitations exist in our study. First, as a cross-sectional study, we are prevented from concluding causal inferences regarding the relationships between SRN connectivity and worry. The sample consists of older adults, thus we cannot generalize our findings to younger populations. We included individuals with varying levels of cognitive function, which may impact the results, as cognitive deficits are known to influence anxiety. We did not measure any markers of hormones so any of those interpretations should be investigated thoroughly in future studies. Finally, the sample is relatively homogenous in terms of education and racial background, primarily consisting of well-educated White individuals, which also limits generalizability of results to more diverse populations.

In conclusion, we presented new data describing the positive association between SRN connectivity and worry in older adults, an association that is strengthened with age despite an overall age-related decline in SRN connectivity. We also found that women showed less of a reliance on emotional suppression for emotion regulation and greater language cognitive function than men and this was associated with greater and more effective top-down regulation of SRN. These findings highlight SRN connectivity as a neural correlate of worry, with the role of ECN-SRN connectivity remaining an important target for future research.

## Data Availability

All data produced in the present study are available upon reasonable request to the authors.

## Acknowledgements

We would like to acknowledge the staff of the ARGO Neuroscience of Aging Research Group for their work and support.

